# Deficit in intersegmental torques drives post-stroke stiff-knee gait

**DOI:** 10.1101/2025.05.19.25327905

**Authors:** Serhii Bahdasariants, Ana Maria F. Barela, Cheryl Brandmeir, Odair Bacca, Sergiy Yakovenko

**Author notes:** Corresponding author: Sergiy Yakovenko, **Email:**. **Competing Interest Statement:** The authors declare no competing interests.

## Abstract

Walking, a seemingly simple task for many, becomes a challenge for people after a stroke. Typically, the swinging leg kinematics is fine-tuned to provide toe clearance, but reduced knee flexion after stroke requires adaptive gait strategies, such as hip hiking and circumduction, to prevent stumbling. The cause of reduced knee flexion in a dynamic system is unintuitive and often obscured by muscle weakness, hyperactivity, or abnormal joint coordination. Using physical models, we predicted that intersegmental forces may be largely responsible for the deficits. Leveraging subject-specific inverse modeling of body dynamics, we compared force generation in post-stroke and neurotypical participants in overground walking. We tested if the contribution of active muscle-generated and passive intersegmental torques from neighboring segments may be responsible for the observed reduced knee flexion. The similar levels of active knee torque across both study groups at the onset of swing indicated that knee interaction torque is the main cause; the component of this torque generated at the hip was decreased (more than that at the ankle) in the post-stroke group. Identified deficit in hip flexion torque at swing onset unveils a key biomechanical mechanism underlying reduced knee flexion post-stroke, providing a physics-informed target for post-stroke gait rehabilitation.

## Introduction

Stroke is a leading cause of adult disability worldwide, affecting millions of individuals who often struggle with walking impairments (Balaban and Tok, 2014; Martin et al., 2025). Among these impairments is stiff-knee gait, characterized by insufficient knee flexion during the swing phase (Fig. 1a) (Li, 2023). Under normal conditions, the knee flexes to facilitate foot clearance and maintain forward momentum, but in hemiparetic stroke, this motion is reduced, forcing survivors to adopt compensatory maneuvers such as hip hiking and circumduction (Fig. 1b) (Kerrigan et al., 2000). These compensations can slow walking speed, diminish endurance, and limit functional mobility. Although stiff-knee gait commonly occurs in stroke survivors, its biomechanical underpinnings remain incompletely understood (Krajewski et al., 2025).

**Fig. 1.**
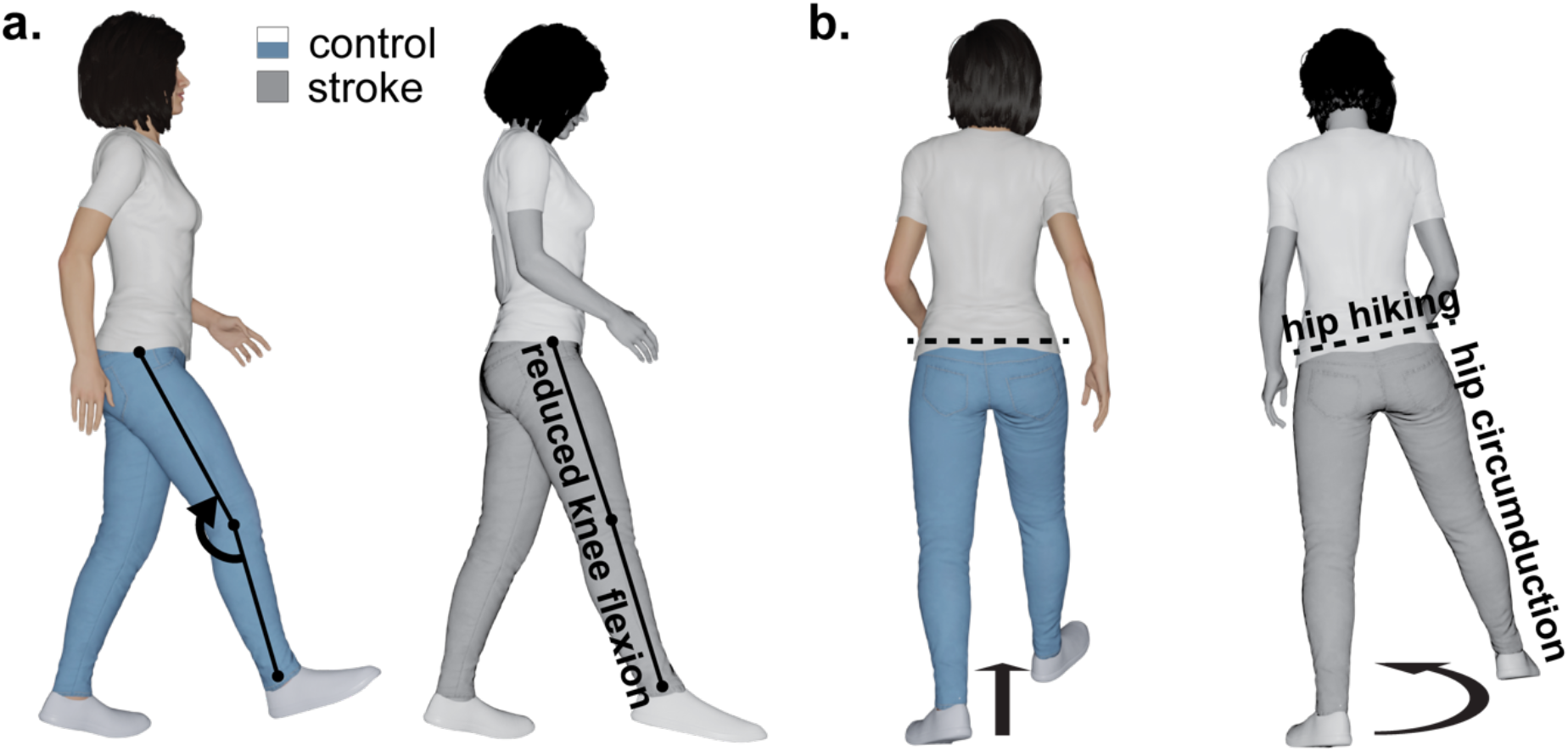
Characteristic gait impairments and compensatory mechanisms post-stroke. (**a**) During the swing phase, stroke survivors (grayscale) often exhibit reduced knee flexion and inadequate ankle control (label omitted for compactness), contrasting with the typical kinematics observed in healthy individuals (color). (**b**) To compensate, common strategies include hip circumduction and hip hiking on the affected side. Created in Blender: Bahdasariants, S. (2025).

In healthy gait, the ankle’s late-stance plantarflexor moment generates forward propulsion and initiates knee flexion (Zajac and Gordon, 1989; Lipfert et al., 2013). One hypothesis proposes that weakened or delayed ankle push-off after stroke reduces knee flexion velocity, contributing to stiff-knee gait (Gatti et al., 2012; Apti et al., 2016). Musculoskeletal models support this premise, showing that diminished ankle power can lower peak knee flexion (Piazza and Delp, 1996; Anderson et al., 2004). Interventions that enhance push-off, such as functional electrical stimulation, can partially—but not fully—restore knee motion (Kesar et al., 2009; Kobayashi et al., 2015; Sekiguchi et al., 2020), indicating that limited ankle power alone does not account for the full spectrum of knee flexion impairments.

During the stance-to-swing transition in unimpaired walking, quadriceps activity subsides, allowing the knee to flex. Another hypothesis attributes stiff-knee gait to prolonged or excessive knee extensor torque, often attributed to rectus femoris overactivity or spasticity (Chantraine et al., 2005; Robertson et al., 2009; Tenniglo et al., 2023). Electromyography findings show that, in some stroke survivors, quadriceps activation persists into swing, inhibiting knee flexion (Waters et al., 1979; Kerrigan et al., 1991). Targeted interventions, including botulinum toxin injections or rectus femoris transfers, can improve knee motion (Gage, 1994; Robertson et al., 2009; Tok et al., 2012; Dreher et al., 2014; Tenniglo et al., 2023). However, not all patients exhibit abnormal knee-extension moments or respond to these treatments, implying that quadriceps overactivity alone does not fully explain stiff-knee gait (Goldberg et al., 2006; Stoquart et al., 2008).

Hip flexors normally accelerate the thigh forward in pre-swing, producing an interaction torque that promotes knee flexion (Piazza and Delp, 1996; Fox and Delp, 2010). A third hypothesis argues that hip flexor weakness or impaired coordination reduces this forward acceleration and, consequently, limits knee flexion (Kerrigan et al., 1999; also see Fujita et al., 2024). Experimental work shows that selective hip flexor fatigue in healthy participants induces a stiff-knee–like pattern (Akalan et al., 2016). Computational models further predict that restoring hip flexor strength can enhance knee flexion (Kerrigan et al., 1998). Nonetheless, some individuals with stiff-knee gait do not exhibit notable hip flexor deficits, indicating the possibility of additional mechanisms (Riley and Kerrigan, 1998).

In this study, we thus addressed the incomplete understanding of stiff-knee gait by conducting a comparative analysis of joint torques in chronic stroke survivors (n=20) and age-matched controls (n=20). Overground walking kinematics were recorded, and a subject-specific inverse dynamic model was applied to decompose torques into muscular, intersegmental, and gravitational components (Bahdasariants et al., 2023). This approach isolates each joint’s contribution to knee flexion during swing, clarifying how mechanical factors combine to produce stiff-knee gait. The findings may inform targeted, patient-specific interventions aimed at restoring safer and more efficient walking.

## Materials & Methods

This study aimed to examine the contribution of active and passive torques to aberrant knee flexion in individuals with stroke. Our methodology involved three steps: (i) collecting populational data on stroke and healthy gait kinematics (Fig. 2a), (ii) conducting biomechanical inverse dynamics simulations to determine net torques (Fig. 2b) and their components (Fig. 2c), which induced experimentally recorded movements, and (iii) identifying torque components correspondent with abnormal gait patterns through cross-populational statistical analysis. Subsequent sections expand on these three steps.

**Fig. 2.**
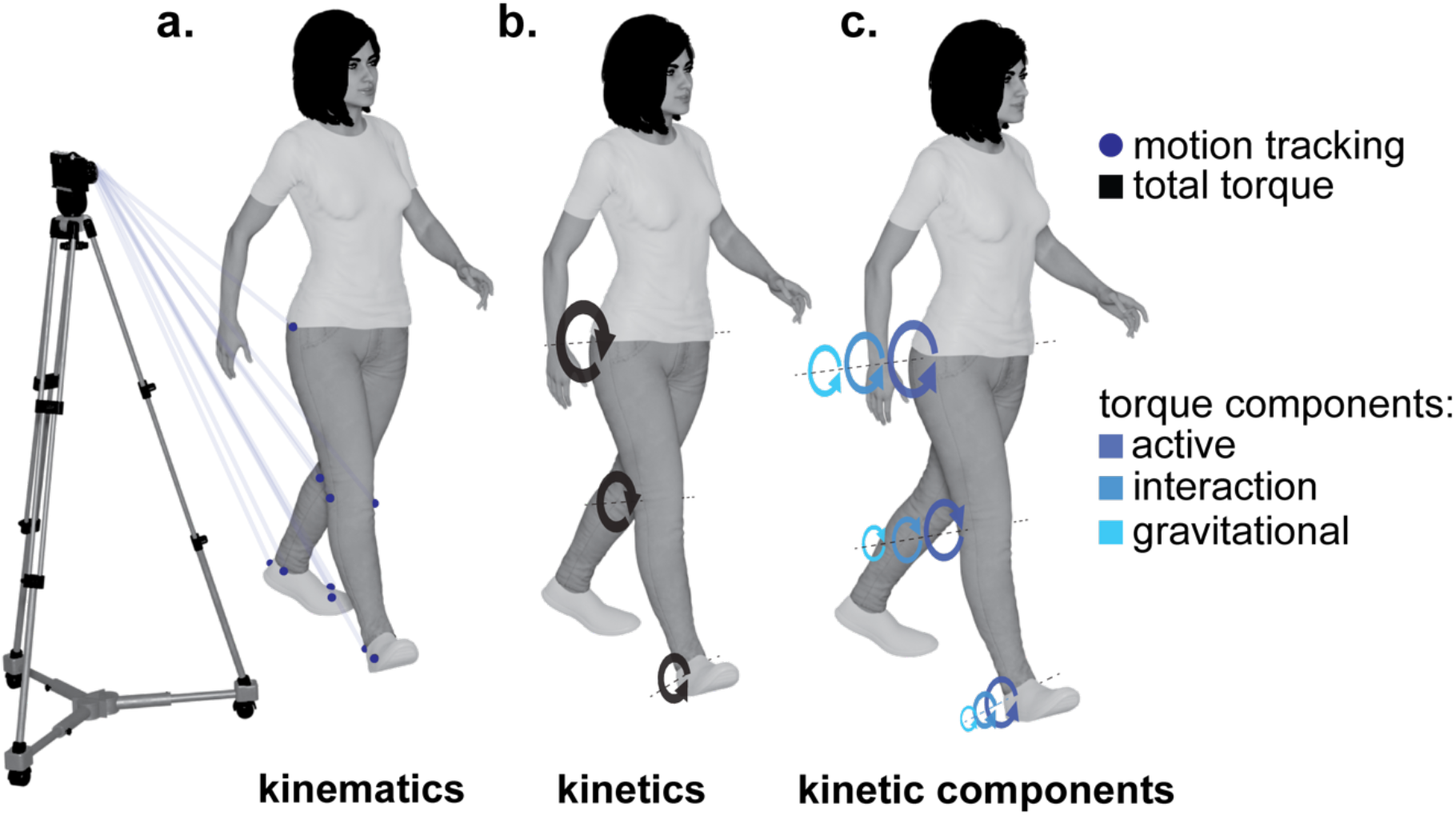
Quantitative data for analyzing post-stroke gait. Panel (**a**) illustrates kinematics, detailing body movements without consideration of underlying forces. Panel (**b**) shows kinetics, specifically total joint torques (represented in black) instigating observed movement. Panel (**c**) demonstrates the total joint torques decomposed into active, interaction, and gravitational components, represented by different shades of gray arranged from darkest to lightest, respectively. Created in Blender: Bahdasariants, S. (2025).

### Experimental kinematics

Three-dimensional marker trajectories were recorded for 20 stroke and 20 neurotypical controls. The two groups were matched by age and sex (see Table 1 for demographics). Ethical clearance was obtained from the Ethical Review Committee of Cruzeiro do Sul University, and participants provided written informed consent (ethical review committee CAAE 02887518.2.0000.8084). A motion capture system (Vicon, Oxford Metrics, Inc.) consisting of eight infrared cameras was used to track reflective markers placed on anatomical landmarks, following the guidelines of the International Society of Biomechanics (Wu et al., 2002). These included the anterior superior iliac spine, posterior superior iliac spine, medial and lateral epicondyles of the femur, tibial tuberosity, medial and lateral malleoli, and first and fifth metatarsals. Before data collection, a calibration trial was conducted with participants standing in a T-pose to align marker clusters with anatomical landmarks. The markers were then removed from the medial and lateral femoral epicondyles, tibial tuberosity, and medial and lateral malleoli. All participants were instructed to walk naturally at a comfortable pace on a ten-meter walkway equipped with two force platforms (Kistler, Model 9286BA). Adequate time was allotted for participants to practice and familiarize themselves with the experimental and laboratory setting before the recording session. The experimental recordings yielded a total of 300 swing phases for stroke participants and 303 swing phases for neurotypical controls, detected using foot-strike and toe-off events, which, in turn, were identified from the ground reaction forces and vertical velocities of the feet (O’Connor et al., 2007). Joint angles for the hip, knee, and ankle, along with the standing foot rotation relative to the ground, were derived from the marker trajectories using the biomechanics analysis software Visual3D (C-Motion, Inc.). Computed angles were low-pass filtered with a 2^nd^-order Butterworth filter (cutoff frequency of 10 Hz) to remove motion artifacts and minimize noise interference (Winter, 2009 p.65). Subsequently, the angles were resampled at 200 Hz using cubic splines to improve the accuracy of biomechanical simulations performed with the fixed-step implicit Euler method.

**Table 1.**
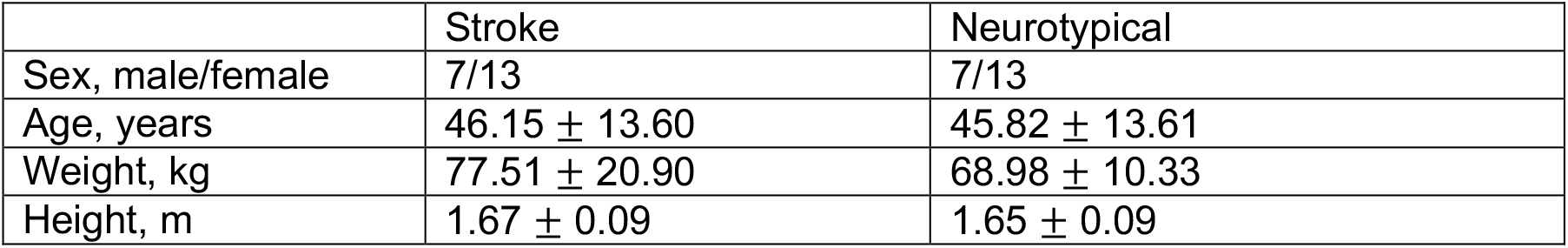
Key demographic characteristics of the studied populations. The age, weight, and height values are presented in mean ± standard deviation format.

|  | Stroke | Neurotypical |
| --- | --- | --- |
| Sex, male/female | 7/13 | 7/13 |
| Age, years | 46.15 ± 13.60 | 45.82 ± 13.61 |
| Weight, kg | 77.51 ± 20.90 | 68.98 ± 10.33 |
| Height, m | 1.67 ± 0.09 | 1.65 ± 0.09 |

### Simulated kinetics

Joint torques inducing the experimentally recorded motion were extracted using inverse dynamics simulations. These torques encompassed the active (τ_*act*_), interaction (τ_*int*_), and gravitational (τ_*g*_) components interconnected as follows:

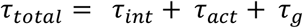

Here, the total torque (τ_*total*_)represents the combined force required to rotate a joint at a specific angle, considering external forces and the effects of joint interactions. The force exerted on a joint due to the movement of its neighboring joints is defined as interaction torque (τ_*int*_). This interaction torque becomes noticeable during activities like walking, where the movement of one joint, for instance, the hip, influences the torque experienced by another joint, such as the knee. The active torque (τ_*act*_) represents the muscle-induced force required to rotate a joint in the absence of external forces (τ_*g*_ = 0) and joint interactions (τ_*int*_ = 0). To grasp the nature of this torque, imagine an astronaut moving in a weightless environment (τ_*g*_ = 0). If the astronaut’s hip and ankle are immobilized and no interaction forces are transmitted to the knee (τ_*int*_ = 0), the sole torque component capable of inducing knee movement is the active torque (τ_*act*_). Lastly, the gravitational component of torque (τ_*g*_) is the torque working against the gravitational force.

The extraction of torque components involved five steps. First, the total torque (τ_*total*_) was determined by solving the inverse dynamics problem using the implicit Euler method, with an integration step inversely proportional to the sampling frequency (Δ*t* = 5ms). Secondly, the inverse dynamics was solved again, though without accounting for gravity (τ_*g*_ = 0), producing driving torques of a combined value of active and interaction components (τ_*act*_ + τ_*int*_). To separate these two components, the third step focused on simulating the active torque for each joint. This was accomplished by nullifying the gravity force (τ_*g*_ = 0) and constraining the angular excursion of all but one joint at a time. The procedure was performed independently for the ankle, knee, and hip joints, yielding the approximate values of active torque (τ_*act*_) for each joint. In the fourth step, the obtained values of active torque (τ_*act*_) were subtracted from the sum of active and interaction torques (τ_*act*_ + τ_*int*_) calculated in the second step. This subtraction allowed for estimation of the interaction torque component (τ_*int*_). Finally, in the fifth step, the gravitational torque component was computed by subtracting the active and interaction torques from the total torque calculated in the first step (τ_*g*_ = τ_*total*_ − τ_*act*_ − τ_*int*_). All computed torque components were used to assess kinetic differences between stroke and normal gait swing phase patterns.

### Biomechanical model

The simulation of torques during the swing phase was conducted using our previously developed Simscape Multibody (R2023a, MathWorks) model of the lower body detailed elsewhere (Bahdasariants et al., 2023). Briefly, the leg segments were represented by solid frustums of right circular cones, with their inertial properties, centers of mass, and joint locations estimated using Hanavan’s formulations (Hanavan, 1964). Proximal base diameters of the thigh and shank were calculated based on sex- and age-specific anthropometric tables (McDowell et al., 2005; Fryar et al., 2021), while the foot segment diameter matched the malleolus height. Distal base diameters were estimated through regression equations (Hanavan, 1964), and segments’ axial lengths were derived from Winter’s tables (Winter, 2009 p.86). Motion between segments was described using rotational primitives. We originally modeled the knee with a one-dimensional rotational primitive, and the hip and ankle with three-dimensional rotational primitives. To replicate the foot center of pressure during the single stance phase (Fuchioka et al., 2015), the standing foot was connected to the ground through a three-dimensional rotational primitive around the estimated anatomical position of the metatarsophalangeal joint. In this paper, we focused on the DOFs most relevant for forward propulsion of the lower extremity during the swing phase, thus, we only actuated flexion-extension DOFs in the hip, knee, and ankle of both legs, as well as the elevation of the standing foot. This reduced the model complexity to seven actuated DOFs, while the remaining DOFs were not considered and remained immobile.

### Statistical Analysis

The Kolmogorov-Smirnov test was used to examine the normality of the joint angle and torque distributions during the initial swing phase, as well as the distribution of swing phase durations. The Kruskal-Wallis test was employed to compare the non-normal angle and torque distributions between the stroke and healthy groups. Fisher’s least significant difference procedure was used for the multiple comparison test. A one-sample t-test was used to compare the non-normal swing phase duration distributions. All tests were conducted using the Statistics and Machine Learning Toolbox (MATLAB R2023a, MathWorks) with significance level set to 5%.

## Results

A total of 40 volunteers participated in this study: 20 chronic stroke survivors (7 males, 13 females) and 20 age-matched neurotypical controls (7 males, 13 females); see Table 1 for demographic details. We first verified the presence of knee flexion impairments during swing in the stroke group by measuring peak-to-peak angular excursions of the knee. On average, the stroke group, walking significantly slower (Fig. 3d), exhibited a 1.75-fold decrease in peak-to-peak angular excursions of the knee compared to controls (Fig. 3a, middle panel), confirming the deficit. Closer inspection revealed that, although knee postures at the start of swing were not significantly different between groups (Fig. 3b, middle boxplots), the stroke group consistently showed reduced knee excursion toward terminal swing (Fig. 3c). This progressive divergence in knee motion motivated our analysis of potential deficits at the ankle, knee, or hip that might drive the observed discrepancy.

**Fig. 3.**
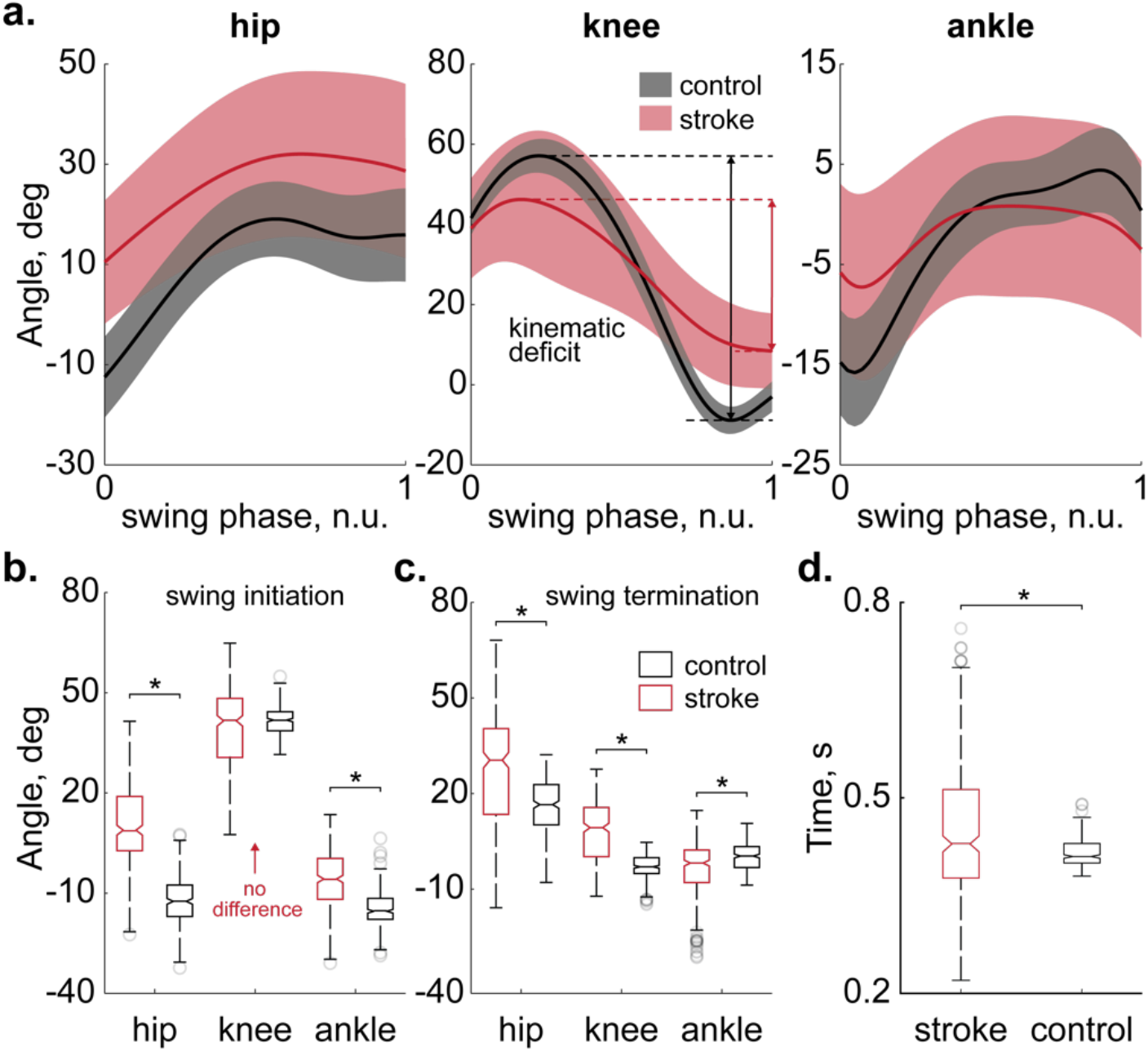
Swinging leg kinematics in stroke and control groups. (**a**) Average joint angles during the swing phase (solid lines) are shown for the hip, knee, and ankle joints in twenty stroke (n=303 swing phases, red) and twenty control (n=300 swing phases, black) participants. The shaded areas represent one standard deviation. Peak-to-peak amplitudes of knee excursions are shown for stroke (red arrow) and control (black arrow) participants using horizontal double-sided arrows. (**b**) Distributions of joint angles measured at the onset of the swing phase reveal significant differences (p<0.01, denoted by asterisks) in the hip and ankle joint excursions, while no significant difference is observed for the knee joint excursions between stroke (red boxes) and control (black boxes) groups. Horizontal solid lines represent the medians, while the bottom and top edges of the box indicate the 25th and 75th percentiles, respectively. Whiskers extend to the extremes, and gray circles depict outliers. The description of the boxplots in **c** and **d** is consistent with **b**. (**c**) Distributions of joint angles measured at the end of the swing phase show significant differences across all joints (p<0.01). (**d**) The swing phase durations differed significantly between the groups (p<0.01).

First, we considered the possibility that reduced ankle push-off during late stance could yield inadequate momentum for knee flexion in swing. This mechanism predicts diminished active torque at the ankle and, consequently, lower interaction torque transmitted to the knee. When we decomposed net joint torques into active muscular, gravitational, and intersegmental (interaction) components, we observed significant post-stroke reductions in active ankle torque compared to controls: approximately a 65% decrease (see second column of Fig. 4c for qualitative comparisons and the same column of Fig. 5c for quantitative comparisons). However, although this deficit was statistically significant (p<0.01, Kruskal-Wallis), the absolute torque reduction (0.0212 Nm/weight/height) was relatively small. First columns of Fig-s 4c and 5c likewise indicate that the ankle’s contribution to swing-phase dynamics was less dramatically altered than that of the hip (see below). These findings suggest that, while weakened ankle push-off was present, it alone did not account for the marked reductions in knee excursion.

**Fig. 4.**
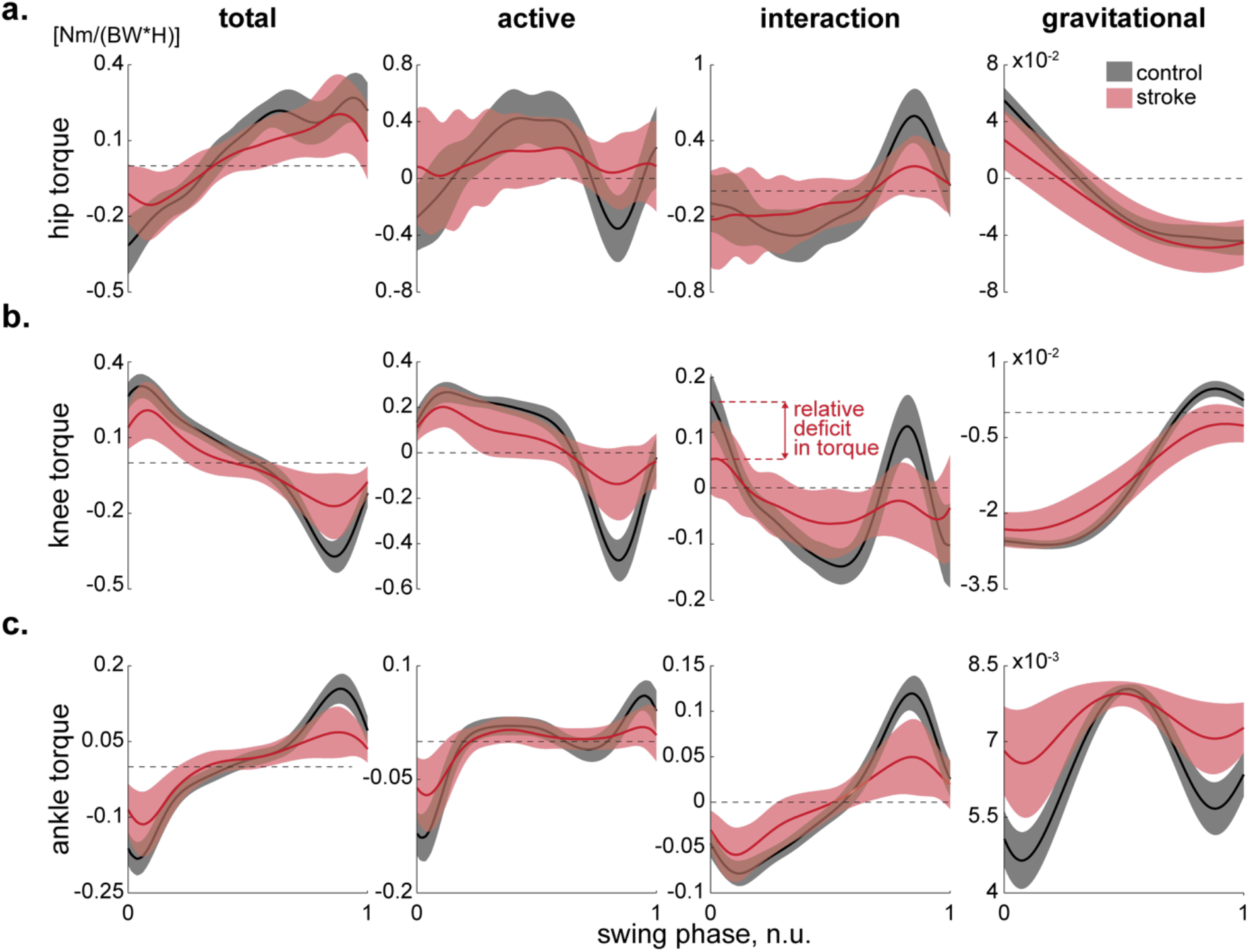
Swinging leg kinetics in stroke and control groups. Average joint torques (solid lines) of the hip (**a**), knee (**b**), and ankle (**c**) joints, normalized to subjects’ body weight and height, are shown for twenty stroke (n=303 swing phases, red) and twenty healthy (n=300 swing phases, black) participants. The shaded areas represent one standard deviation. Total, active, interaction, and gravitational torques of hip are shown on Panel **a**. The total torque reflects the force required to move the joint at the experimentally recorded angle, considering external and interaction forces. The active torque represents muscle force needed to rotate the joint to the desired angle when no external or interaction forces are applied. The interaction torque originates from other joints, and the gravitational torque opposes gravity. Notably, differences between total and active torques in the hip are prominent. The description of Panels **b** and **c** are similar to Panel **a** with one key exception. In Panel **b**, active and gravitational torques exhibit similarities between populations, while the interaction torque is decreased (indicated by a double-sided red arrow), suggesting its primary role in reducing the total knee torque. Abbreviations: BW—body weight and H—height.

**Fig. 5.**
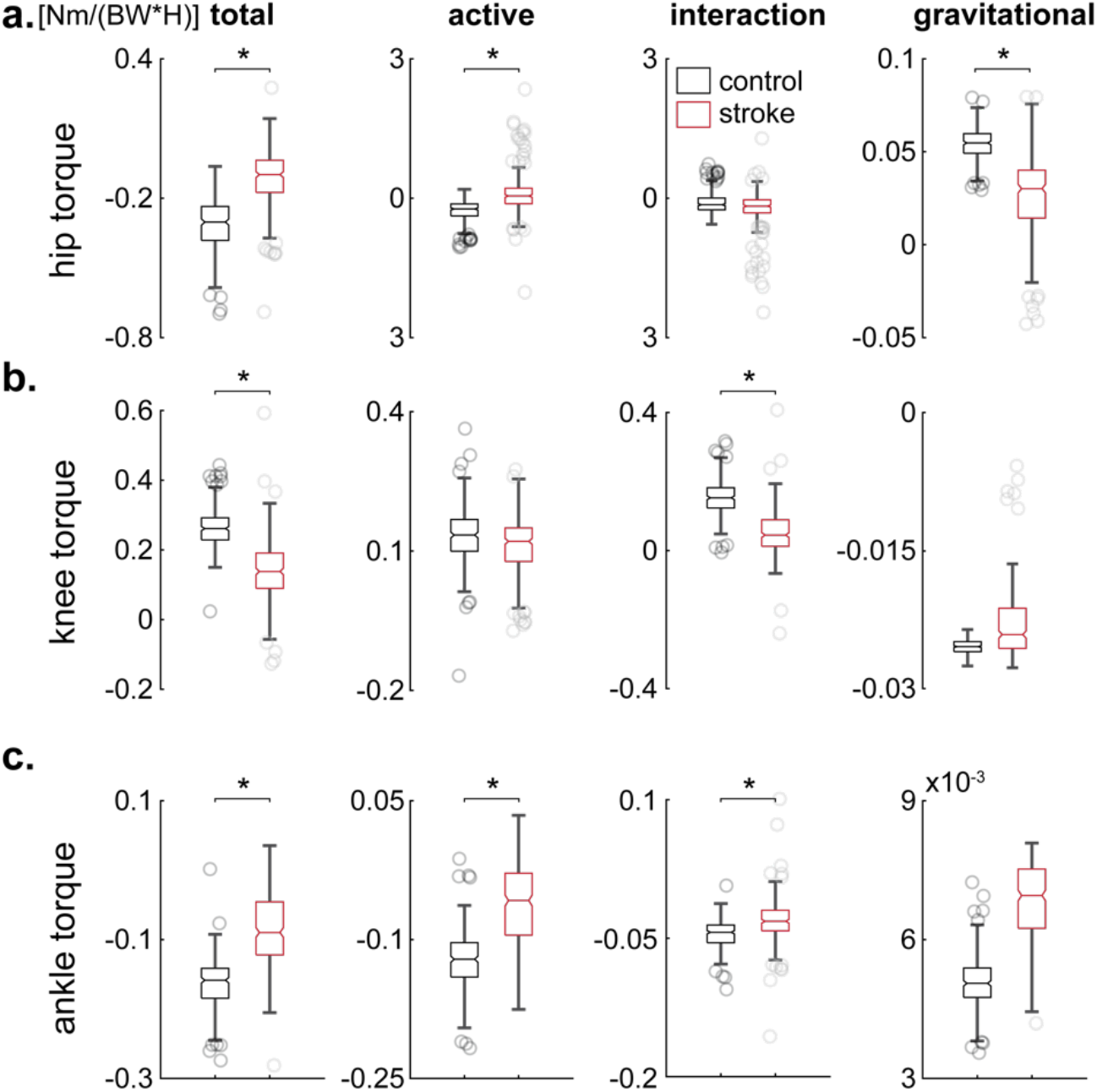
Distributions of the normalized torques at the onset of the swing phase. (**a**) Values of total, active, interaction, and gravitational hip torques measured at the beginning of the swing phase are plotted for twenty stroke (n=303 swing phases, red) and twenty healthy (n=300 swing phases, black) participants. Significant differences in total, active, and gravitational torques are observed (p<0.01, denoted by asterisks). (**b**) The difference in total knee torque arises from a deficient interaction torque propagating from the hip and ankle to the knee. Active and gravitational torques show no significant differences between the stroke and control groups. (**c**) Similar to Panel **a**, the decrease in total ankle torque is attributed to a deficient active torque, yet the interactions from the knee were expectedly reduced, and gravitational torques remained similar to those observed in the control group. The boxplots’ description is the same as in Fig. 3B. Abbreviations: BW— body weight and H—height.

Next, we tested the idea that abnormal muscle activation at the knee (e.g., excessive knee extensor activity) underlies stiff-knee gait. Under this scenario, we would expect substantial changes in active knee torque that deviate from controls. Yet our inverse dynamics analysis indicated that active knee torque was similar between the stroke and control groups at the beginning of the swing (see second column of Fig. 4b for qualitative comparisons and the same column of Fig. 5b for quantitative comparisons). The passive (interaction and gravitational) torques around the knee, however, were altered (see third and fourth columns of Figs. 4b and 5b), indicating a significant shift in how the knee was being driven by torques generated elsewhere. This pattern is inconsistent with a simple overactivity isolated to the knee—if knee muscles were the primary contributors, we would expect to see a pronounced surplus in the active knee torque itself. Instead, the principal changes arose from how torques transferred among the joints, suggesting that the cause of the knee impairment was likely more proximal or distal.

Given the modest role of the ankle and the negligible differences in active knee torque, we next evaluated whether insufficient hip flexor power might explain the observed reduction in knee excursion. In normal gait, a robust burst of hip flexor activity during late stance and early swing accelerates the thigh forward, thereby generating intersegmental torques that flex the knee (Piazza and Delp, 1996). We found that stroke survivors exhibited a 52% decrease in active hip torque relative to controls (p<0.01, Kruskal-Wallis; see the second column of Fig. 5a), corresponding to a deficit of 0.2033 Nm/weight/height—nearly ten times larger than the ankle torque drop (0.0212 Nm/weight/height). Meanwhile, interaction torque at the hip (third column in Fig. 5a)—which partially reflects contributions from the trunk and other proximal segments—*did not differ* between groups, indicating that stroke survivors did not significantly modify trunk or pelvis mechanics to compensate for their diminished hip strength. Although gravitational torque at the hip (fourth column in Fig. 5a) shifted, this likely reflects minor postural adjustments intended to offload or stabilize the weakened hip. Thus, the net deficit in hip torque primarily arose from a lack of active flexor power, reducing the momentum imparted to the knee and thereby limiting its swing-phase excursion.

In sum, none of the three commonly hypothesized single-joint mechanisms—excessive knee extensor activity, insufficient ankle push-off, or inadequate hip flexor power—fully accounted for the pronounced reduction in swing-phase knee excursion among our stroke cohort. Rather, our findings point to a combination of impairments, with the *shortfall in active hip flexor torque exerting the most substantial impact* and weakened ankle push-off also contributing to a lesser degree. Consequently, although knee overactivity was not a primary driver of stiff-knee gait in these participants, both proximal (hip) and distal (ankle) deficits interacted to limit knee flexion during swing.

## Discussion

The primary finding of this study is that among post-stroke ambulators with stiff-knee gait, impaired hip flexor function was the largest contributor to reduced swing-phase knee flexion (Fig. 6). In our cohort, weakness or insufficient activation of the hip flexors corresponded to markedly diminished knee flexion during swing, whereas distal ankle push-off power played a secondary role. This contrasts with the traditional view that attributed stiff-knee gait mainly to excessive quadriceps (rectus femoris) activity (Kerrigan et al., 1991; Chantraine et al., 2005; Stoquart et al., 2008; Akbas et al., 2020). Instead, our results align with evidence that weak hip flexors can directly lead to knee hyperextension in mid-stance and inadequate knee flexion torque in swing. Consistent with this mechanism, simulation analyses of hemiparetic gait have shown reduced iliopsoas (hip flexor) contributions to leg swing initiation in individuals with limited knee flexion (Brough et al., 2022). Notably, we found no support for the notion that knee extensor overactivity was the major limiting factor for swing-phase knee excursion in this cohort (Fujita et al., 2020), even though all participants exhibited the defining reduced knee flexion angle of stiff-knee gait post-stroke.

**Fig. 6.**
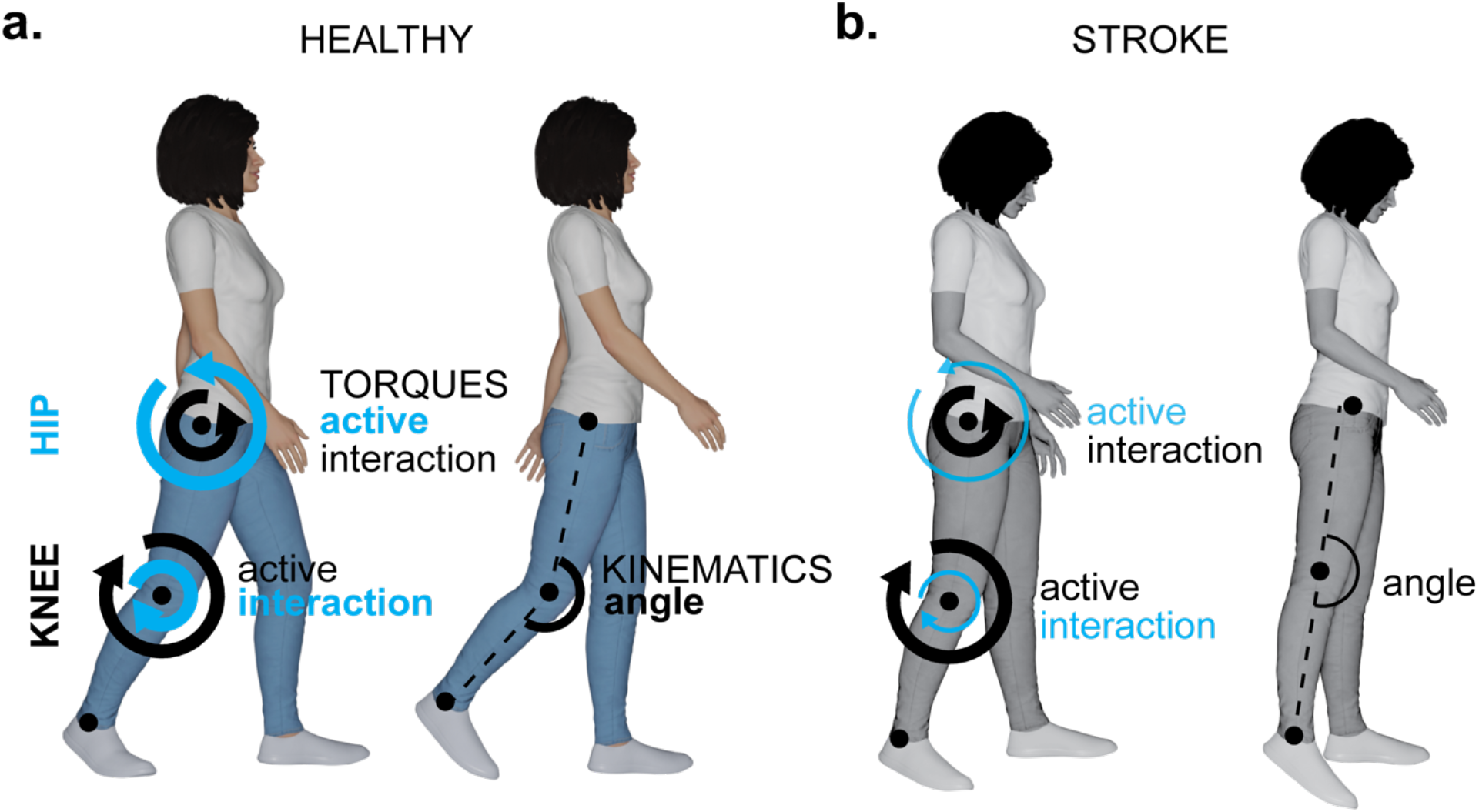
Reduced intersegmental interaction forces originating predominantly from the hip, and to a lesser extent the ankle, result in diminished knee flexion following stroke. In able-bodied participants (**a**), the combination of active and interaction torques at the hip, supplemented by those from the ankle, generates a propulsive moment essential for normal limb advancement, facilitating sufficient knee flexion and foot clearance. In stroke survivors (**b**), substantially reduced active torque production at the hip, accompanied by minor reductions at the ankle, leads to decreased intersegmental interaction forces at the knee. This reduction markedly compromises knee flexion amplitude during swing, compelling patients to adopt compensatory strategies for foot clearance. Created in Blender: Bahdasariants, S. (2025).

A secondary contributing factor to stiff-knee gait in these participants was insufficient ankle plantarflexor push-off. Consistent with the general reduction in paretic propulsion observed after stroke, inadequate push-off in late stance can diminish the knee’s flexion velocity at toe-off, leading to a smaller swing-phase knee flexion angle. For example, Campanini et al. (2013) reported that about 85% of post-stroke stiff-knee cases were explained by deficient push-off, with knee flexion appearing as a “bottom-up” response to ankle power generation. Moreover, interventions that augment plantarflexor output during pre-swing have demonstrated improvements in knee kinematics: functional electrical stimulation of the plantarflexors in late stance significantly increases peak knee flexion in swing (Kesar et al., 2009). Our findings corroborate these observations by confirming that reduced ankle push-off contributes to swing knee-flexion deficits, although to a lesser extent than hip flexor weakness. It is important to note, however, that push-off deficits do not fully account for the impairment in all cases. Total propulsive force has not always predicted swing-phase knee flexion in prior studies (Campanini et al., 2013), and abnormally high braking impulses (due to premature co-activation or insufficient propulsion) have been linked with reduced knee flexion and compensatory circumduction during swing (Dean et al., 2020). This suggests that while ankle plantarflexor weakness is an important factor, stiff-knee gait is usually multi-factorial, and proximal hip power plays a more dominant role when push-off is especially diminished.

Notably, our analysis did not implicate abnormal knee extensor activity (e.g. spastic or prolonged rectus femoris activation) as a primary mechanism for the limited knee flexion. Early work had identified excessive quadriceps activity as a feature of stiff-legged gait in spastic paresis (Waters et al., 1979; Kerrigan et al., 1991), and clinicians often assess rectus femoris overactivity in post-stroke gait evaluations (Yelnik et al., 1999). However, accumulating evidence indicates that quadriceps hyperactivity is not the chief driver of stiff-knee gait for many patients. Consistent with our results, a recent simulation study found that the rectus femoris contributed minimally to impeding knee flexion in most post-stroke individuals, whereas the knee extensors acting in late stance (vasti muscles) had a larger braking effect associated with lower knee flexion angles (Brough et al., 2022). Our cohort’s lack of significant rectus femoris influence is in line with Campanini et al. (2013), who noted that only 15% of their patients showed stiff-knee gait caused by a knee “braking” (extensor overactivity) mechanism. The modest benefits observed with treatments targeting the rectus femoris further support the secondary role of knee extensors—for example, blocking or denervating the rectus femoris yields on average only a 7–9° improvement in peak knee flexion during swing (Tenniglo et al., 2014). This degree of change, while statistically significant, is relatively small (also see Stoquart et al., 2008 for non-responders), and helps explain the limited functional impact of interventions solely aimed at reducing quadriceps spasticity. Together, these findings reinforce that, in the absence of unusually high extensor hyperactivity, stiff-knee gait after stroke is driven more by inadequate generation of propulsion and swing initiation than by active knee-extension resistance.

These insights have important clinical and biomechanical implications. Rehabilitation strategies for post-stroke stiff-knee gait should prioritize strengthening and augmenting the paretic hip flexors and ankle plantarflexors to address the identified primary deficits. For instance, targeted hip flexor strengthening and high-intensity gait training that enhances ankle push-off power may yield significant improvements in swing-phase knee excursion. Notably, improved plantarflexor function has been shown to increase knee flexion velocity and foot clearance during gait (Knarr et al., 2013; Fujita et al., 2020), and stronger hip flexor engagement can compensate for distal weakness to restore more normal knee motion (Brough et al., 2022). As such, treatments focused solely on mitigating knee extensor spasticity (such as botulinum toxin injections or surgical rectus femoris release) should be reserved for the subset of patients who demonstrate clear evidence of quadriceps overactivity contributing to the gait impairment. Our findings, in line with prior reports, suggest that many patients will experience only minimal knee kinematic gains from rectus femoris injections if hip and ankle power deficits are not concurrently addressed (Campanini et al., 2013; Tenniglo et al., 2014). Yet, while these findings present a clear direction for targeted interventions, it is also important to acknowledge the limitations of our study that may influence the generalizability and applicability of these results.

A primary limitation of this study is that we did not examine the stance phase mechanics, which constitute about 60% of the gait cycle. Despite this limitation, analyzing the swing phase alone is particularly relevant for addressing common issues seen post-stroke, such as reduced walking speed, increased fall risk, and poor toe clearance (Moore et al., 1993). Another limitation is the use of a model that only considers flexion-extension DOFs. While this simplifies the analysis, it disregards movements in other DOFs, such as rotational and translational movements. However, although these movements contribute to maintaining balance and adapting to uneven surfaces (Winter, 1995), they are not the primary drivers of forward propulsion. Given our focus on the deficient movement of the knee joint in the sagittal plane, we specifically considered DOFs contributing to this motion. Next, a model that uses a revolute joint, which is fixed to the ground, presents both a limitation and a benefit when it comes to estimating experimental ground reaction forces post-stroke. On one hand, this simplified representation of the foot-ground interaction forces might decrease the realism of the simulation; on the other hand, it provides a straightforward computational method for estimating ground reaction forces. This is particularly beneficial when working with participants affected by stroke whose mobility impairments make the experimental recordings challenging. Another important consideration to note is the exclusion of spasticity from the analysis conducted in this study. The focus was primarily on stroke survivors who exhibited mild symptoms (Fig. 7) and possessed the ability to walk independently without the assistance of robotic devices or braces.

**Fig. 7.**
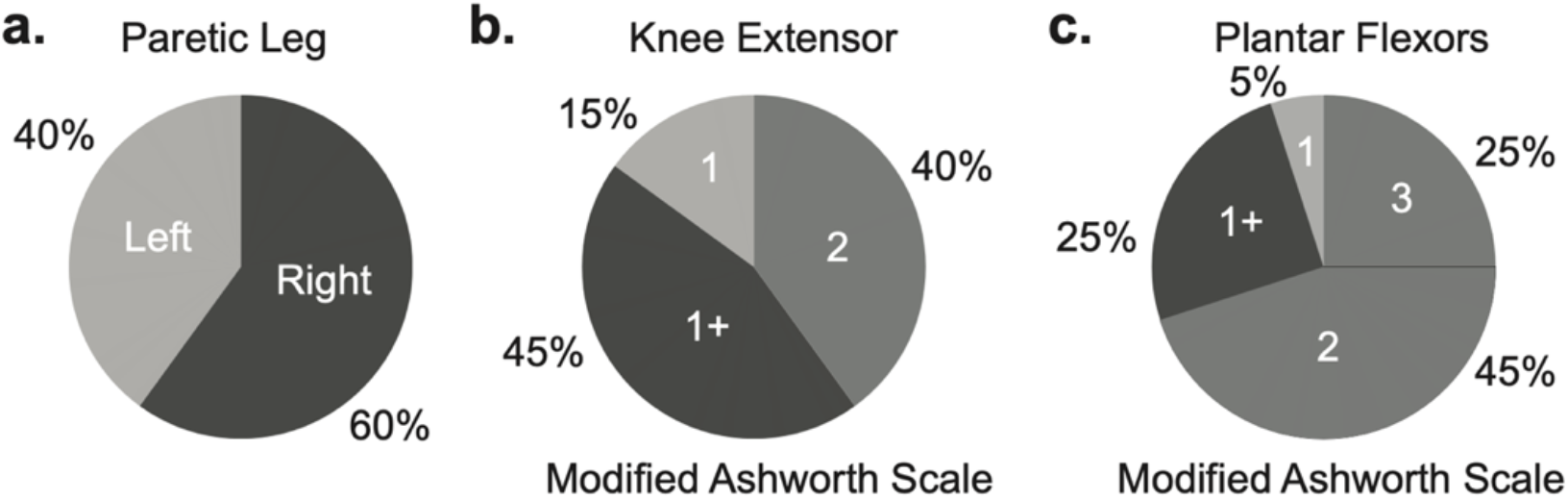
Percentage of patients with left versus right paretic leg (**a**) and their scores on the Modified Ashworth Scale (**b-c**), which illustrate the severity of spasticity.

Consequently, the validity of the current model in situations involving a greater degree of spasticity or when spasticity significantly limits the range of motion remains uncertain. It is unknown whether strength in adjacent joints can effectively counteract the effects of spasticity.

## Conclusions

In this study, we found that hip flexor weakness predominantly limits swing-phase knee motion in post-stroke gait, with insufficient ankle push-off acting as a secondary factor. These findings suggest that focusing rehabilitation efforts on strengthening proximal and distal muscles simultaneously may yield better outcomes than interventions targeting knee extensors alone. While our cohort was limited to independently ambulatory chronic stroke survivors, the observed patterns underscore the importance of tailored rehabilitation programs that consider individual joint deficits. Future work should investigate whether similar results hold for acute stroke populations or those with more pronounced spasticity. By clarifying the biomechanical underpinnings of stiff-knee gait, our results provide a rationale for targeted therapies that enhance overall mobility and reduce fall risk in post-stroke individuals.

## Data Availability

All data produced in the present study are available upon reasonable request to the authors.

## Acknowledgements

This research was funded by the NIH Award 1 R03 HD099426-01A1 and the São Paulo State Research Foundation (FAPESP) Grant 2018/04964-8. Additional support was provided by the West Virginia University Center for Foundational Neuroscience Research and Education, the West Virginia University Stroke and Its Alzheimer’s Disease Related Dementias (ADRD) T32 Program, the West Virginia Foundation Distinguished Doctoral Scholarship Program, the Mitochondria, Metabolism & Bioenergetics (MMB) Working Group, and the Community Foundation for the Ohio Valley Whipkey Trust.

